# Re-contacting biobank participants: lessons from a pilot study within FinnGen

**DOI:** 10.1101/2022.04.07.22273501

**Authors:** Rodosthenis S. Rodosthenous, Mari E.K. Niemi, Lila Kallio, Merja Perälä, Perttu Terho, Theresa Knopp, Eero Punkka, Enni M. Makkonen, Paula Nurmi, Johanna Mäkelä, Pauli Wihuri, Marco Hautalahti, Corianna Moffatt, Paolo Martini, Laura Germine, Viola A. Mäkelä, Oona A. Karhunen, Jari Lahti, Tero S. Hiekkalinna, Tero Jyrhämä, Huei-Yi Shen, FinnGen, Heiko Runz, Aarno Palotie, Markus Perola, Andrea Ganna

## Abstract

**Background:** Biobank studies are well suited to identify novel disease risk factors. However, these studies often offer limited insights into participants’ lifestyle, behavioral, or cognitive characteristics, among others. FinnGen is a nation-wide biobank project in Finland and one of the largest in the world that has successfully linked genetic and register-based information for 259,578 individuals (N=500,000 by 2023); however, it lacks cognitive, behavioral, and lifestyle measurements.

**Methods:** We conducted a pilot study to evaluate the feasibility of obtaining cognitive, behavioral, and lifestyle information by re-contacting 5,995 participants from three FinnGen-participating Biobanks. We created an online portal to allow participants to securely log-in, provide consent, and access the study survey tools.

**Results:** The overall participation rate was 18.6% (23.1% among individuals aged 18-69). A second reminder letter yielded an additional 9.7% participation rate in those who did not respond to the first invitation. Re-contacting participants via an online healthcare portal yielded lower participation than re-contacting via physical letter. The completion rate of questionnaire and cognitive tests was high (92% and 85%, respectively), and measurements were overall reliable among participants. For example, the correlation (*r*) between self-reported body mass index and that collected by the biobanks was 0.92.

**Conclusions:** In summary, this pilot suggests that re-contacting FinnGen participants with the goal to collect a wide range of cognitive, behavioral and lifestyle information without additional engagement, results in a low participation rate, but with reliable data. We suggest that such information be collected at enrollment, if possible, rather than via post-hoc re-contacting.

## Introduction

Biobank studies are being set up across the world^1-9^. These studies are characterized by the possibility to link biological measurements, such as DNA, proteins, and metabolites with extensive longitudinal health information. Health outcomes are often obtained by linkage with electronic health records or national registers, as in the case of the Nordic countries. Biobank studies have allowed extensive characterization of the genetic architecture of common diseases^10-12^, provided novel epidemiological insights^13 14^ and identified novel disease markers^15^.

The UK Biobank study^1 16^, one of the largest and most widely used biobanks, has also collected lifestyle, behavioral information, and anthropometric measurements for all their participants. This was possible, at the cost of collecting a non-representative sample of the population^17-19^, because all participants were enrolled via in-person visit to one of 22 the recruitment centers. However, most of the other biobank studies use a different approach collecting samples via the healthcare system or via other approaches that do not entail an extensive in-person examination at recruitment. For this reason, it is often difficult to obtain extensive behavioral and lifestyle information from biobank participants and re-contacting after recruitment is required.

The FinnGen study is a public-private partnership research project combining genotype data generated from Finnish biobank samples and digital health record data from Finnish health registers (https://www.finngen.fi/en) aiming to provide new insight in disease genetics^20^. Up to 500,000 participants will be part of FinnGen and > 350,000 have already been genotyped and linked with comprehensive health registers. Participants to the FinnGen study are recruited by several biobanks across Finland and all participants have signed a broad Biobank consent in accordance with the Finnish Biobank Act. Participants are enrolled because they are part of previous research studies or via hospitals and blood donation centers, but no extensive behavioral and lifestyle information is systematically collected for everyone at recruitment.

Overall, FinnGen does not suffer from the “healthy volunteer effect” and, on the contrary, is enriched for individuals that are more likely to have been in contact with the healthcare system. Samples from consented individuals can be used across many research projects, if approved by the biobanks. Thus, individuals are not actively informed of their participation in FinnGen. Here we describe the results of a pilot study that aimed at re-contacting FinnGen participants with the goal to collect cognitive, behavioral and lifestyle information via a custom-made online platform.

## Results

In this genetically informed pilot study, we invited 5995 FinnGen participants across three Biobanks who were selected according to the inclusion criteria shown in **Figure 1**. Out of 5995 individuals invited, 1115 individuals accessed the study online portal and successfully completed the general questionnaire with an overall participation rate of 18.6% **(Table 1)**. The participation rate in the 18-69 years old age group was slightly higher at 23.1% (1018/4399). The highest overall participation rate across all ages was observed in Helsinki Biobank with 23%, followed by Tampere Biobank with 17% and Auria Biobank with 15.8%. Lower participation rate in Tampere and Auria biobanks is partially explained by the higher age range of the invited individuals. Among the 18-69 years old range, the participation rate was similar across all biobanks. The highest participation rate was consistently observed in the 40-69yo age group across all biobanks **(Table 1 and Supplementary Table 1)**. The participation rates among individuals who were ≥70yo was substantially lower (6.1%) as compared to the <70yo group (23.1%). We also tested if sending a follow-up reminder letter would significantly improve participation rate among non-responders. To assess this, we sent a reminder by post to 422 individuals from Tampere (n=240) and Auria (n=182) Biobanks who were invited initially but did not participate in the study during the first phase. The overall response rate for this second letter was 9.7% (41/422), higher in Tampere Biobank (11.5%, 28/240) than Auria Biobank (7.1%, 13/182).

**Table 1.**
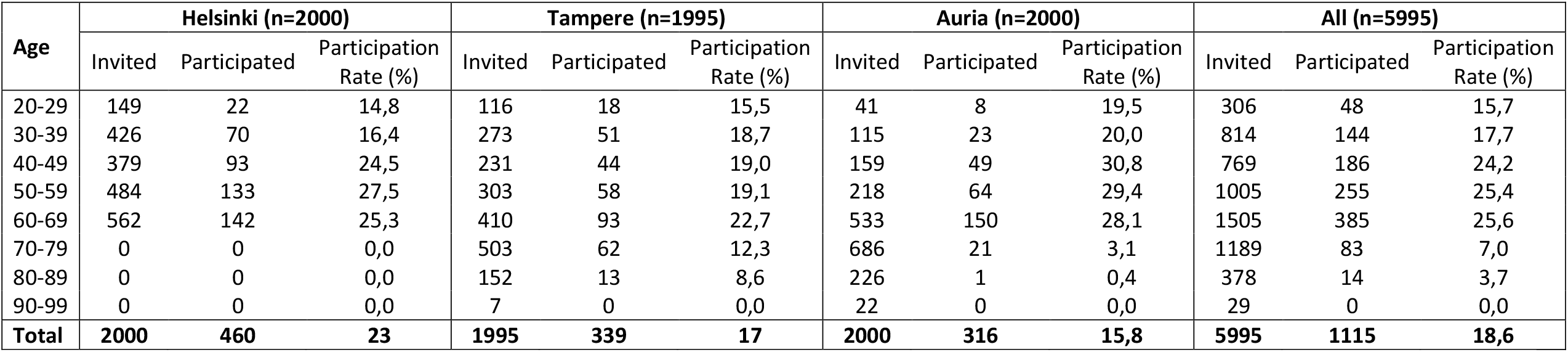
Basic questionnaire participation rates among all invitees by age group and biobank.

| Age | Helsinki (n=2000) |  |  | Tampere (n=1995) |  |  | Auria (n=2000) |  |  | All (n=5995) |  |  |
| --- | --- | --- | --- | --- | --- | --- | --- | --- | --- | --- | --- | --- |
|  | Invited | Participated | Participation Rate (%) | Invited | Participated | Participation Rate (%) | Invited | Participated | Participation Rate (%) | Invited | Participated | Participation Rate (%) |
| 20-29 | 149 | 22 | 14,8 | 116 | 18 | 15,5 | 41 | 8 | 19,5 | 306 | 48 | 15,7 |
| 30-39 | 426 | 70 | 16,4 | 273 | 51 | 18,7 | 115 | 23 | 20,0 | 814 | 144 | 17,7 |
| 40-49 | 379 | 93 | 24,5 | 231 | 44 | 19,0 | 159 | 49 | 30,8 | 769 | 186 | 24,2 |
| 50-59 | 484 | 133 | 27,5 | 303 | 58 | 19,1 | 218 | 64 | 29,4 | 1005 | 255 | 25,4 |
| 60-69 | 562 | 142 | 25,3 | 410 | 93 | 22,7 | 533 | 150 | 28,1 | 1505 | 385 | 25,6 |
| 70-79 | 0 | 0 | 0,0 | 503 | 62 | 12,3 | 686 | 21 | 3,1 | 1189 | 83 | 7,0 |
| 80-89 | 0 | 0 | 0,0 | 152 | 13 | 8,6 | 226 | 1 | 0,4 | 378 | 14 | 3,7 |
| 90-99 | 0 | 0 | 0,0 | 7 | 0 | 0,0 | 22 | 0 | 0,0 | 29 | 0 | 0,0 |
| **Total** | **2000** | **460** | **23** | **1995** | **339** | **17** | **2000** | **316** | **15,8** | **5995** | **1115** | **18,6** |

**Figure 1.**
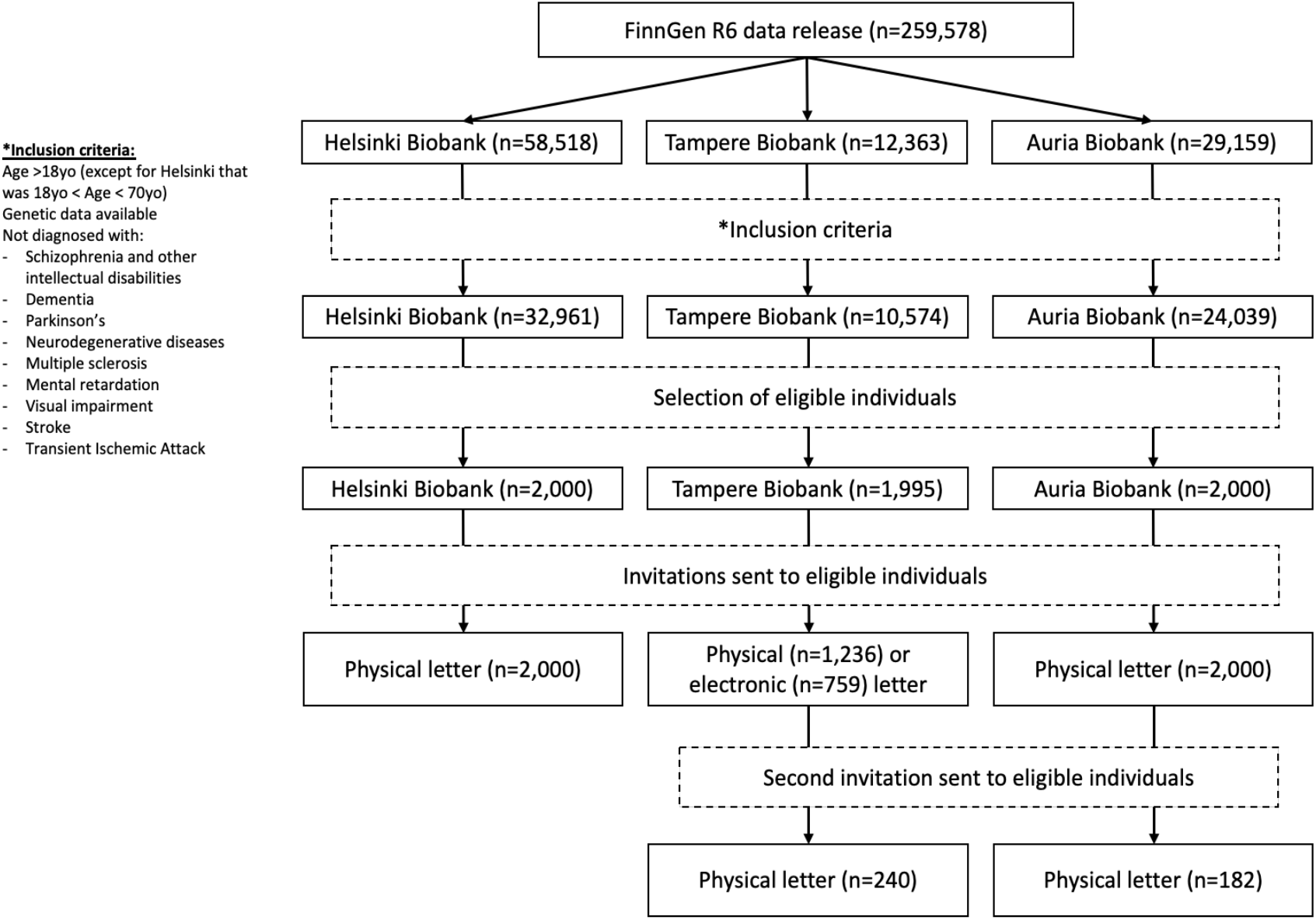
Flow diagram depicting the selection of study participants.

Using FinnGen data, we further investigated whether any differences in basic characteristics and disease prevalence among invitees could have potentially impacted their participation in the study. No significant age difference was observed between invitees who participated (54.9±13.5, [mean±SD]) versus those who did not participate (55.6±15.6, [mean±SD]) in the study (p=0.08); however, we found that invitees who participated had a lower BMI (28.0±6.2, [mean±SD]) compared to those who did not participate (28.5±6.2, [mean±SD]) in the study (p=0.02). We also found that invitees with a university degree or higher were more likely to participate (24%, 433/1810) as compared to invitees who had a lower education (17%, 519/3099). The difference in the two proportions between the groups was statistically significant (p<0.0001). When assessing health information using FinnGen data, we found that a significantly higher (p<0.0001) proportion of invitees who did not participate were previously diagnosed with hypertension (24.9%, 1210/4854) as compared to invitees who participated (16.9%, 186/1101). No differences in other disease prevalences, such as asthma, arthrosis, depression, and immune bowel disease were observed between the two groups.

In addition, we compared the participation rate between individuals invited via physical letters and those invited electronically via the Tampere Biobank OmaTays healthcare portal. Out of 1995 invitees, 759 received their invitations via the OmaTays healthcare portal and 1236 via physical letter. The participation rate was 12.1% among those invited via the OmaTays compared to 21.4% among those who were invited via a physical letter from Tampere Biobank. Data retrieved from the OmaTays healthcare portal showed that 451/759 (59.5%) of those who were invited electronically did not open the invitation at all. Among the rest who viewed the invitation, ∼30% (92/308) went on to participate in the study. Individuals invited in this pilot study were consented in the first place via the Biobank consents, in accordance with the Finnish Biobank Act. Thus, they could decide to withdraw their consent for their samples to be used in research studies. In this study, a 0.3% of the invited individuals contacted any of the three biobanks to withdraw their consent.

Among individuals who started answering the online questionnaire, the completion rate was 92%. The completion rate for the entire battery of cognitive tests was 85%, despite the length of the tests (estimated to be 30-40 min). However, substantially fewer people started the cognitive test compared to the general questionnaire (n=699 vs n=1115). Among 143 participants who provided their experience through the feedback questionnaire, the median NPS score was 8, indicating that most individuals were happy with the current design. More specifically, 121/143 (85%) gave a score >7, which is interpreted as good/very good. On the contrary, 22/143 (15%) scored the questionnaire with a 6 or less, indicating they had a negative experience (**Figure 2**). It is worth mentioning that only 53% (73/137) had previous experience with online questionnaires, whereas for 47% (64/137) of the participants this was their first online questionnaire.

**Figure 2.**
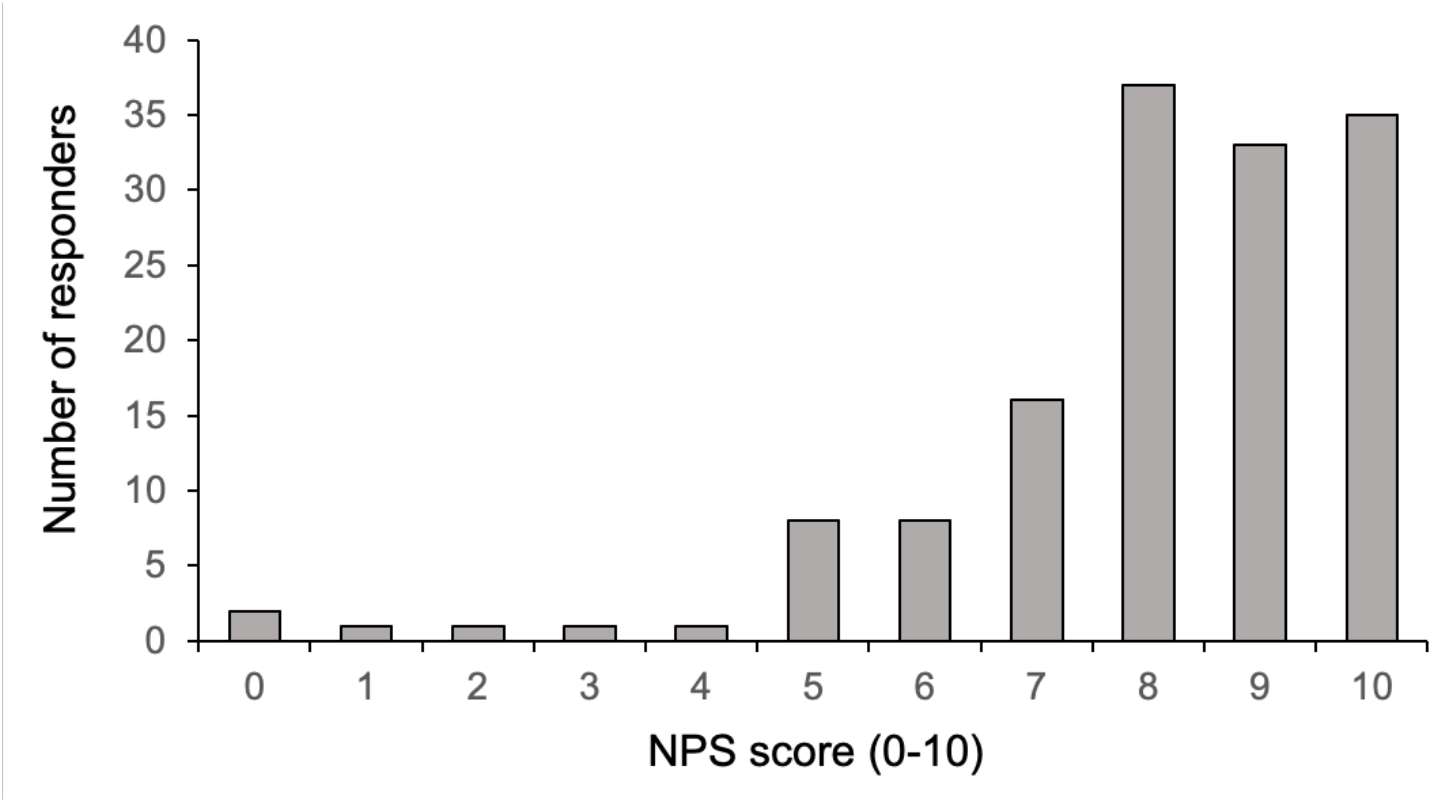
Net Promoter Scores (NPS) from the feedback survey asking for the experience of participants completing the online questionnaires (n=143).

For a subset of FinnGen participants, information about body mass index (BMI) was available and extracted from electronic health records or in-person visits. Thus, for 673 individuals, we could compare the self-reported BMI data obtained from the questionnaire with those previously available in FinnGen and extracted from electronic health records or in-person measurements. We found a high correlation between the two measurements (Spearman’s *r*=0.92) despite these being collected at different time points and with different approaches (**Figure 3**). The mean±SD of the self-reported and FinnGen collected BMI data was 28.1±6.0 and 28.0±6.2, respectively. Similarly, we could compare self-reported disease diagnoses from the questionnaire with disease information available in FinnGen and obtained from national health registers **(Supplementary Table 2)**. In the questionnaire, participants were asked whether they had been professionally diagnosed with any of the listed diseases. Overall, we noticed a higher prevalence for most diseases when self-reported compared to when extracted from the health registers. For example, asthma had a prevalence of 11.3% (124/1101) when data was extracted from the health registers compared to 18.5% (204/1101) when self-reported via the online questionnaire. We observed similar trend with vitiligo: The prevalence of vitiligo from health register data was 0.1% (1/1101) *vs* 1.7% (19/1101) when self-reported. On the other hand, in the case of primary sclerosing cholangitis, we found the same prevalence of 0.7% (8/1101) between the health register and self-reported data. For some diseases, a higher prevalence is expected because health registers do not provide good coverage if diagnosed in primary care. On the other hand, some individuals might over-report or mis-report disease diagnoses. A head-to-head comparison between self-reported and register-based diseases is challenging because different combinations of diagnostic and medication codes can be used to define the same disease from health registers.

**Figure 3.**
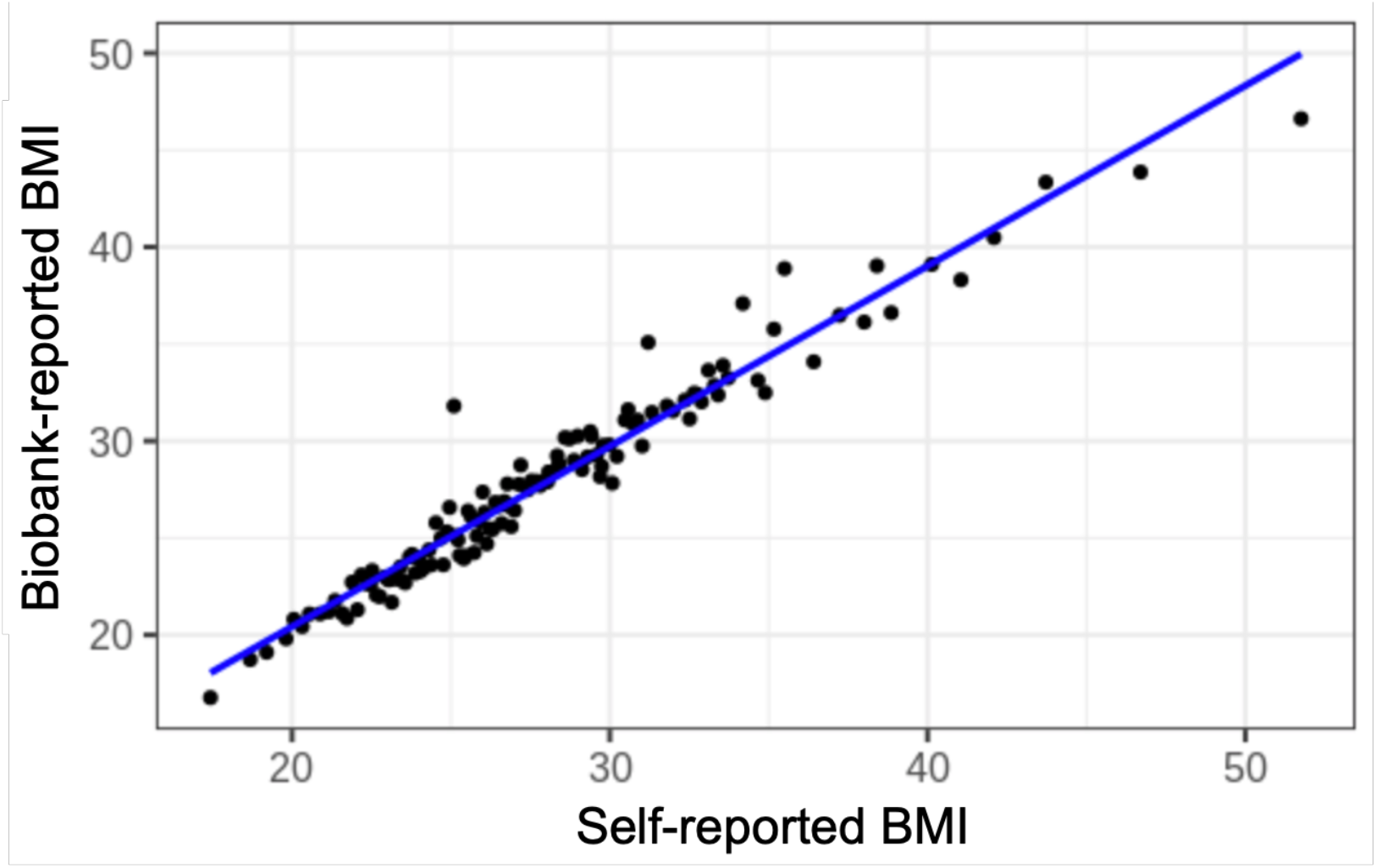
Spearman’s correlation between biobank-reported and self-reported body mass index (BMI) measures acquired from the same participants (n=647).

Last, access to genetic data for both participants and non-participants in this pilot study allowed us to test if our strategy of recruiting individuals in the top and bottom 3% of a polygenic score (PS) for cognitive abilities resulted in differential study participation between the two groups. We found that a higher PS for cognitive performance was associated with higher participation. In particular, 19.4% of individuals who participated in the pilot study were in the top 3% of the PS for cognitive performance compared to 16.9% of those that did not participate. The difference in the two proportions between the groups was statistically significant (p=0.01) but modest **(Figure 4)**.

**Figure 4.**
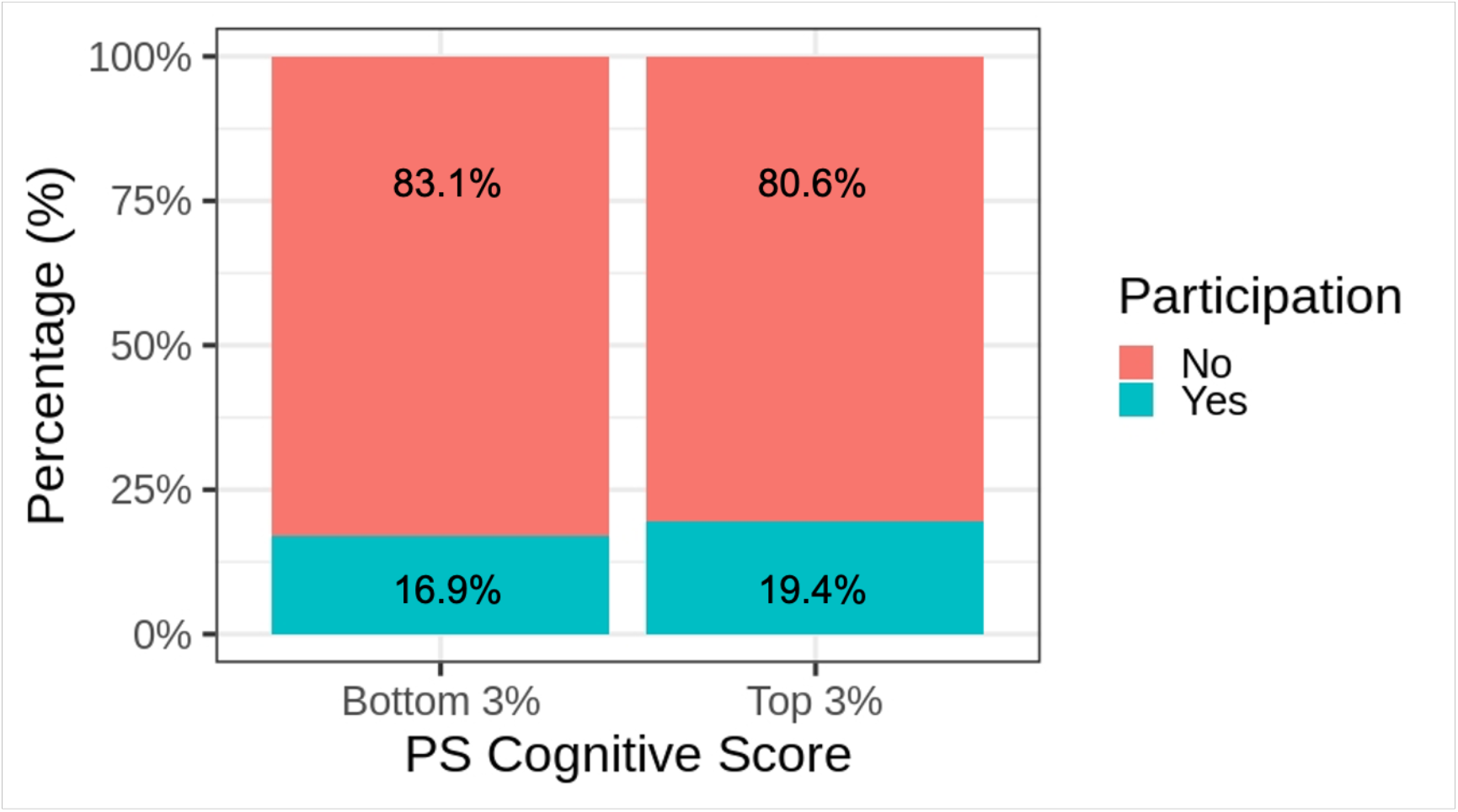
Stacked bar graph showing the participation proportion of study participants by PS cognitive performances group.

## Discussion

Several biobank studies are designed to prioritize scalable and economical sample collection by using existing biological banks or hospital-based recruitment strategies. Often, extensive characterization of lifestyle, cognitive and behavioral information is done *a posteriori* by re-contacting study participants. In this pilot study we have proposed and tested a strategy to re-contact participants in one of the largest biobank studies in the world: FinnGen. To this goal, we have established a scalable re-contacting process and designed an online re-contacting platform (i.e., OmaBiopankki) for secure identification of participants. The platform can now be utilized as a benchmark for future re-contacting studies in Finland.

Despite declining response rates in population-based surveys globally^21^, research studies conducted in Finland have shown a high participation rate (>50%)^22-25^. In this pilot study, we have observed a lower participation rate of 23% in the 18-69 years old age range, which may be explained by several reasons. First, all participants provided a general consent to their biobank that covers current studies such as FinnGen and a wide range of possible future research studies. Therefore, consented individuals may not be aware of their participation or be directly engaged at the time of contacting on behalf of the FinnGen study. Second, this pilot study was designed to capture a broad range of cognitive, behavioral, and lifestyle information from invited individuals and not to target any specific disease(s). The three biobanks included in this pilot recruit individuals that are hospitalized or have been directly in contact with the healthcare system. Thus, studies that target specific diseases directly relevant to each participant’s own health may result in higher participation rates. Third, for the same reason, consenting participants are likely to be sicker than the general population, which may subsequently impact their participation in such studies. In fact, lower participation rates among less healthy individuals are well documented^26 27^.

As part of our efforts to boost the participation rate in this study, we tested several methods. Initially, we restricted the age group of invitees to under 70 years old because we observed a significantly lower participation rate among those over 70 years old. Based on the feedback we got through our study contact email and phone helpline, we believe that this difference was mainly due to the limited access or lack of familiarity with the use of the internet and mobile banking applications required for secure authentication among older individuals. In addition, we sent a follow-up letter as a reminder to a subset of non-responders to the initial invitation and observed a significant boost in response rate (9.7%). Our finding is very comparable to Harrison’s et. al. study that found a 9% increase in participation rates after sending a reminder letter to non-responders^28^. Another study by Smith et.al., also found that sending follow-up letters significantly increases the chance of response among non-responders to the initial invitation^29^. We also evaluated response rates between sending a physical or an electronic letter and found that invitees who received a physical letter were much more likely to respond to the survey. In a recent study, researchers tested the same invitation methods and found a striking difference in participation rates when sending a physical (26.8%) or an online (1.8%) invitation^28^.

To the best of our knowledge, this is one of the first studies that assessed participation rate in relation to the cognitive PS score of participants. To further support our finding that individuals with a higher cognitive PS score are more likely to participate in online surveys, we also compared the education level between those who participated compared to those who did not participate in the study. We found that invitees who had a university degree or higher were more likely to participate (24%, 433/1810) as compared to invitees who had a lower education (17%, 519/3099).

In conclusion, this pilot suggests that re-contacting individuals that have consented to be part of a biobank study with the goal to collect a wide range of cognitive, behavioral and lifestyle information can be challenging and may result in lower-than-expected participation. We speculate that returning some tangible incentive and/or relevant health information to participants might improve participation rates. Future studies are warranted to test this hypothesis and other strategies to improve participation in such survey research studies. Nonetheless, we suggest that cognitive, behavioral and lifestyle information be collected, whenever possible, at enrollment rather than via a post-hoc re-contacting process.

## Methods

### Study population

We used data from FinnGen release R6, which included 259,578 individuals with genetic information available. According to the Finnish Biobank Act (688/2012), biobanks may re-contact a person who has given such permission in his/her biobank consent. Three biobanks (Helsinki, Auria and Tampere) encompassing 100,040 FinnGen participants (Helsinki n=58,518; Auria n=29,159; Tampere n=12,363) participated in the pilot study. We included individuals that were 18 years or older at the time of the initiation of the study (Feb 2021). For Helsinki biobank, we restricted inclusion to individuals younger than 70 years of age. We excluded individuals with severe neuropsychiatric disease, or cognitive or physical disabilities, as we expected the participation rate to be lower in this group. More specifically, we excluded individuals with any of the following diagnoses as obtained from the health register data: 1) Any dementia (ICD-10: G30,F051,F00-03); 2) Neurodegenerative diseases (ICD-10: G310-312, G318-319); 3) Parkinson’s disease (ICD-10: G20) 4); Multiple sclerosis (ICD-10: G35); 5) Schizophrenia, schizotypal and delusional disorders (ICD-10: F20); 6) Mental retardation (ICD-10: F70-73, F79-78); 7) Stroke (ICD-10: I60-64); 8) Transient ischemic attack (ICD-10: G45); 9) Visual impairment including blindness (binocular or monocular) (ICD-10: H54). Finally, we selected individuals in the top and bottom 3% of a polygenic score (PS) for cognitive performances^30^. This was motivated by previous work showing a strong association between a PS for cognitive performances and study participation^19 31^. Our aim was to prospectively assess if recruiting individuals based on this PS would result in a different study participation.

### Invitation procedures

Invitation was carried out via postal letter for 5995 FinnGen participants by Helsinki (n=2000), Auria (n=2000) and Tampere (n=1236) biobanks. For an additional 759 individuals from Tampere biobank, an invitation was sent electronically via the OmaTays healthcare portal, which has been used already for medical communication and online booking appointments by the Tampere biobank. A random subset of 422 participants aged 18-69 from Tampere (n=240) and Auria (n=182) biobank that did not reply to the first invitation were re-contacted with a second invitation letter.

### Portal for enrollment and collecting survey information

We collaborated with the Finnish biobank cooperative (FINBB) to create a portal that allows study participants to securely log-in, identify themselves, provide consent, and access the study survey tools. The portal is hosted at https://omabiopankki.fingenious.fi/ and secure authentication is guaranteed by a bank ID identification system which is available to most Finnish residents. Once the participants log into the system, the study invitation and consent can be viewed online. After the participants consent to participate in the study, they can view the two information collection tools: a general questionnaire and a battery of the cognitive tests. By selecting each tool, participants firstly view a short description prior to their completion. Both the general questionnaire and cognitive tests are developed on third-party platforms. Once the participants have completed the online questionnaire or cognitive test battery, they are redirected back to the Omabiopankki portal where they can complete the remaining tasks.

### General study questionnaire

The general study questionnaire included 18 categories of questions which cover a broad range of health-related topics that are not obtainable or are difficult to obtain through hospital and register data. All questions were accompanied by the option “don’t know” and “prefer not to answer” to allow the participants to voluntarily skip any of the questions. The following categories were included, with the scientific justification: 1) General questions (sex at birth, height, weight); 2) Women’s health; 3) Smoking; 4) Family medical history; 5) Disease diagnoses (including diseases not well captured by hospital and register data); 6) Medication history (including over-the-counter medications); 7) Alcohol consumption; 8) Early onset neurodevelopmental disorders and psychiatric disorders; 9) Mental health and mood; 10) Education levels; 11) Physical activity; 12) Multi-site pain; 13) Risk taking; 14) Flu and viral infections (respiratory); 15) Sleep; 16) Oral health; 17) Diet; 18) Sauna habits.

### Cognitive tests

We collected data from a test battery designed to capture different aspects of cognitive abilities. This test battery was provided by *TestMyBrain* and translated into Finnish. *TestMyBrain* has previously validated all their cognitive test batteries [https://psyarxiv.com/dcszr/]. Eight different tests were performed: Digit Symbol Matching, Flicker Change Detection, Visual Paired Associates, Multiracial Emotion Identification, Gradual Onset Continuous Performance, Matrix Reasoning, and Vocabulary test. Because the vocabulary test could not be directly translated from English, we created a Finnish-specific version of the test by selecting 19 out of 30 candidate words that provided with the highest correlation with the well-established Wechsler Adult Intelligence Scale -Revised (WAIS-R) Vocabulary task^32^ in a sample of N=24 Finns (79.2% women) with an average age of 24.3 years (SD: 3.0). Our new online vocabulary test had a Spearman’s correlation of 0.86 with the Vocabulary task from the WAIS-R. The analysis of collected cognitive test data is out of the scope of this paper and therefore not shown here. Participants could see a summary of their results and how they scored in each test compared to the average scores after completion of all the tests.

### Feedback questionnaire

To get a sense of the participants’ experience upon recontacting, we introduced a feedback questionnaire directed to the individuals logging into the Omabiopankki portal. The main question to participants was to rate their overall experience in completing the online questionnaire and cognitive tests using a Net Promoter Score (NPS) scale from 0 to 10. In addition, we asked if they have participated in an online questionnaire study before, if they experienced any technical issues while completing the survey, and to share with us what they enjoyed or did not enjoy while completing the survey.

### Ethics

Patients and control subjects in FinnGen provided informed consent for biobank research, based on the Finnish Biobank Act. Alternatively, older research cohorts, collected prior the start of FinnGen (in August 2017), were collected based on study-specific consents and later transferred to the Finnish biobanks after approval by Fimea, the National Supervisory Authority for Welfare and Health. Recruitment protocols followed the biobank protocols approved by Fimea. The Coordinating Ethics Committee of the Hospital District of Helsinki and Uusimaa (HUS) approved the FinnGen study protocol Nr HUS/990/2017. The FinnGen study is approved by Finnish Institute for Health and Welfare (permit numbers: THL/2031/6.02.00/2017, THL/1101/5.05.00/2017, THL/341/6.02.00/2018, THL/2222/6.02.00/2018, THL/283/6.02.00/2019, THL/1721/5.05.00/2019, THL/1524/5.05.00/2020, and THL/2364/14.02/2020), Digital and population data service agency (permit numbers: VRK43431/2017-3, VRK/6909/2018-3, VRK/4415/2019-3), the Social Insurance Institution (permit numbers: KELA 58/522/2017, KELA 131/522/2018, KELA 70/522/2019, KELA 98/522/2019, KELA 138/522/2019, KELA 2/522/2020, KELA 16/522/2020 and Statistics Finland (permit numbers: TK-53-1041-17 and TK-53-90-20). The Biobank Access Decisions for FinnGen samples and data utilized in FinnGen Data Freeze 6 include: THL Biobank BB2017_55, BB2017_111, BB2018_19, BB_2018_34, BB_2018_67, BB2018_71, BB2019_7, BB2019_8, BB2019_26, BB2020_1, Finnish Red Cross Blood Service Biobank 7.12.2017, Helsinki Biobank HUS/359/2017, Auria Biobank AB17-5154, Biobank Borealis of Northern Finland_2017_1013, Biobank of Eastern Finland 1186/2018, Finnish Clinical Biobank Tampere MH0004, Central Finland Biobank 1-2017, and Terveystalo Biobank STB 2018001.

## Supporting information

Rodosthenous_manuscript_MedRxiv_Supplemental_Material.xlsx

Rodosthenous_manuscript_MedRxiv_FinnGen_banner_Authors.xlsx

## Acknowledgements

We want to acknowledge the participants and investigators of FinnGen study. The following biobanks are acknowledged for delivering biobank samples to FinnGen: Auria Biobank (www.auria.fi/biopankki), THL Biobank (www.thl.fi/biobank), Helsinki Biobank (www.helsinginbiopankki.fi), Biobank Borealis of Northern Finland (https://www.ppshp.fi/Tutkimus-ja-opetus/Biopankki/Pages/Biobank-Borealis-briefly-in-English.aspx), Finnish Clinical Biobank Tampere (www.tays.fi/en-US/Research_and_development/Finnish_Clinical_Biobank_Tampere), Biobank of Eastern Finland (www.ita-suomenbiopankki.fi/en), Central Finland Biobank (www.ksshp.fi/fi-FI/Potilaalle/Biopankki), Finnish Red Cross Blood Service Biobank (www.veripalvelu.fi/verenluovutus/biopankkitoiminta) and Terveystalo Biobank (www.terveystalo.com/fi/Yritystietoa/Terveystalo-Biopankki/Biopankki/). All Finnish Biobanks are members of BBMRI.fi infrastructure (www.bbmri.fi). The Finnish Biobank Cooperative - FINBB (https://finbb.fi/) is the coordinator of BBMRI-ERIC operations in Finland. The Finnish biobank data can be accessed through the Fingenious^®^ services (https://site.fingenious.fi/en/) managed by FINBB.

## Data availability statement

Data are available on reasonable request.

## Author contributions

All authors met the criteria of authorship as outlined in the ICMJE and approved this manuscript for submission. Specific contributions were as follows: conceptualisation: AG, MEKN, RSR, MP, AP, JL; methodology: AG, MEKN, RSR, PW, MH, CM, PM, LG, VAM, OAK; project administration and investigation: RSR, LK, MP, PT, TK, EP, EMM, PN, JM, MP, HYS; data curation: MEKN, RSR, PT, PN, TK, RSR, TSH, TJ; formal analysis: RSR, MEKN, AG; writing original draft: RSR, AG; writing, review and editing: JL, HR, AP, MP, AG.

## Supplementary Material

**Supplementary Table 1**. Basic questionnaire participation rates among all invitees under 70 years old by age group and biobank.

**Supplementary Table 2**. Prevalence of the medical history attainment for the same participants (n=1101) between the Re-contacting pilot and FinnGen studies.

